# Who Supports the Caregivers? Perspectives on Mental Health Screening in Paediatrics

**DOI:** 10.64898/2026.06.04.26354967

**Authors:** Nadia Coscini, Rebecca Giallo, Anneke C Grobler, Harriet Hiscock, Melissa Mulraney, Nicole Pope

## Abstract

**Objectives:** To explore caregiver and clinicians perspectives on implementing mental health conversations and supports for caregivers of children with chronic conditions in paediatric outpatient clinics. Specifically, views were sought on (a) screening approaches and measures (phase 1) and (b) how feedback and support could be provided to caregivers experiencing mental health difficulties (phase 2).

**Methods:** Caregivers and clinicians from two outpatient clinics (neuromuscular and diabetes) at a tertiary paediatric hospital in Melbourne, Australia participated in online focus groups in July and August 2024. Caregivers were recruited from outpatient clinics and clinicians were recruited via email. Both groups were combined for phase 1 before separating into breakout rooms for phase 2. Two authors conducted reflexive thematic analysis of transcripts using NVivo.

**Results:** Sixteen participants (caregivers n = 8; and clinicians n = 8) took part in in two semi-structured focus groups. Analysis generated two overarching domains, each comprising multiple themes. Domain 1, “Addressing caregiver mental health,” captured themes of overwhelm and invisibility, diverse caregiving roles, and the need for time and resources to support wellbeing conversations. Domain 2, “Housing the mental health conversation,” encompassed themes of screening preferences, caregiver agency in confidentiality, delivery of feedback, and access to tailored supports.

**Conclusions:** Caregivers and clinicians support routine caregiver mental health discussions in paediatric outpatient settings. Caregivers favour screening at diagnosis and key transitions, with clear, and actionable feedback delivered away from the child. Questions about record-keeping warrant further exploration, as do the perspectives of fathers.

**Article Summary:** This study explored parents’/caregivers’ and clinicians’ perspectives on mental health screening and support for parents of children with chronic conditions accessing paediatric outpatient clinics

**What’s known on this subject:** Parents/caregivers of children with chronic conditions have a high mental health burden, with researchers calling for routine mental health screening in paediatric healthcare settings. Despite growing recognition of this, there is limited data on the implementation of screening methods and referral pathways.

**What this study adds:** Direct consultation with caregivers and clinicians demonstrating they support mental health conversations and being embedded in routine paediatric care. It also demonstrates that caregivers need agency in screening preferences, feedback delivery, confidentiality in records and useful support services.

**Contributors Statement Page:** Dr Nadia Coscini conceptualized and designed the study, designed the data collection instruments, collected data, carried out the initial analyses, drafted the initial manuscript, and critically reviewed and revised the manuscript.

Associate Professor Melissa Mulraney and Dr Anneke Grobler conceptualized and designed the study, supervised data collection and critically reviewed and revised the manuscript.

Professor Rebecca Giallo and Professor Harriet Hiscock conceptualized and designed the study, facilitated the focus groups, supervised data collection and critically reviewed and revised the manuscript.

Dr Nicole Pope led the qualitative analysis including debriefing sessions and iterative theme development, drafted the initial manuscript, and critically reviewed and revised the manuscript.

All authors approved the final manuscript as submitted and agree to be accountable for all aspects of the work.

## Introduction

Chronic conditions (CC) are defined as long-term illnesses that have ongoing complications.^1^ Paediatric CCs^2^ are common, affecting 45% of children (between 3-17 years of age) in the US^3^ and 43% in Australia.^4^ The impact on parents (hence referred to as “caregivers”) is enormous, with effects on their own physical health, family support, career and finances.^5,6^ Caregivers of children with CCs have an increased risk of higher stress and mental health difficulties than caregivers of children without a chronic illness.^7,8^ Meta-analyses found that 35% of caregivers of children with a CC met criteria for depression, 57% for anxiety^9^ and 18.9% for post-traumatic stress disorder.^10^ Caregiver mental health directly affects children’s long-term health outcomes^7,11^ and quality of life,^12^ and is therefore criticla to address.

Integrating caregiver mental health screening into outpatient clinics may provide a practical pathway to support, yet few studies have implemented this approach.^11,13–21^ Our previous systematic review found a paucity of evidence on how best to screen and support caregivers experiencing mental health difficulties.^22^ The largest study, the TIDES study (The International Depression/Anxiety Epidemiological Study) screened 6088 individuals with cystic fibrosis (CF) as well as 4102 caregivers on their mental health, ^11^ informed international consensus guidelines on mental health screening.^23^ However, none of the studies included in the systematic review reported caregiver involvement in the development of the screening programs that were later evaluated, and caregiver input for the CF guidelines occurred only at the guideline development stage.^23^ With recent calls for caregiver mental health screening as a standard of care across all paediatric conditions,^24^ it is essential to involve caregivers in identifying preferred screening tools, delivery methods, communication of results and appropriate support pathways.

The aim of this study was to explore caregiver and clinician perspectives about implementing mental health conversations and supports for caregivers of children with CCs. Specifically, they were asked about (a) current approaches to asking caregivers about their mental health, and (b) their perspectives on potential screening approaches and how caregivers can be provided with feedback and support for mental health concerns during routine outpatient appointments.

## Methods

The study was approved by the institutional review board (HREC 116190) and reported as per the Consolidated Criteria for Reporting Qualitative Research (COREQ).^27^ Prior to being enrolled, participants provided electronic informed consent through REDCap (Research Electronic Data Capture).^28^

### Participants, recruitment and setting

This study employed a qualitative exploratory design^29^ underpinned by a constructivist worldview,^29^ to explore how caregivers and clinicians understand and experience caregiver mental health within paediatric outpatient care. Caregivers and clinicians were purposively recruited from the Royal Children’s Hospital in Melbourne, Australia. Caregivers were eligible if they (a) were the primary caregiver of a child with a CC attending outpatient clinic (diabetes or neuromuscular) aged 0-18 years, and (b) spoke and read English to a level of providing informed consent. Clinicians (e.g., physicians, nurses, allied health staff) were eligible if they provided care to children in these specific outpatient clinics. Caregivers attending clinic with their child between July and August 2024 received a letter informing them of the study and that they may be approached in the clinic waiting room to participate. Clinicians were emailed to request their interest in participating.

### Data collection

Data were collected through semi-structured online focus groups. The interview guide (Supplementary file 1) was developed by the research team, drawing on the biopsychosocial model^25^, and informed by the findings of a systematic review on screening caregivers of children with chronic conditions about their mental health.^22^ The guide included open-ended questions and prompts and allowed flexibility for participants to share their experiences while ensuring core topics were addressed.

Focus groups were conducted in two phases. First, caregivers and clinicians participated together to discuss screening approaches and review example measures (Kessler-10 [K-10] and Depression Anxiety Stress Scales–21 [DASS-21]). Caregivers rom one clinic were grouped with clinicians from the other clinic to minimise pre-existing relationships and support open discussion. Next participants were separated into caregiver and clinician breakout groups to explore perspectives on feedback, support pathways, and preferred modes of communication and enabled both shared discussion and group-specific reflection. Each session included a lead facilitator and supporting facilitators for breakout discussions. Facilitators were experienced researchers in paediatrics, psychology, and qualitative research.

### Data Analysis

We conducted reflexive thematic analysis, following Braun and Clarke’s approach^30^ using to an iterative and interpretive process of familiarisation, coding, and development of themes. Analysis was primarily inductive, with deductive interpretation informed by relevant frameworks and considered both explicit (manifest) and underlying (latent) meanings across full transcripts.^31^

Two authors (NP and NC) led the analysis. Both independently reviewed transcripts and generated initial codes. NP, a clinician–researcher with expertise in qualitative methods, paediatric pain, and family caregiving, brought an emic perspective shaped by clinical practice. NC, as clinician-researcher with expertise in child health and development, brought a complementary perspective. Both engaged in reflexive practices, including memo writing and critical dialogue, to consider how their perspectives shaped interpretation. Themes were developed iteratively through discussion within the wider research team, with representative quotations used to ground interpretations in participants’ accounts.

## Results

Focus groups involved 16 participants (caregivers n = 8; clinicians n = 8) (Figure 1). Eleven eligible clinicians were invited to participate, with 10 signing a consent form, 2 needed to withdraw from focus group 2 due to illness and date unavailability (see supplementary Figure 3). All caregivers were female (n=8, 100%) and clinicians were mostly female (n=6, 75%).(Table 1). Figure 2 introduces a visual illustration of the two domains. Tables 2 and 3 elaborate on the themes within each.

**Table 1.** Demographics of Caregivers and Clinicians attending clinics.

| Demographic | Value |
| --- | --- |
| **Caregivers** | **n = 8** |
| Child age, years: mean (range) | 10.6 (8-14) n=7 |
| Caregiver age, years: mean (range) | 39.3 (39-49) n=6 |
| Gender: female, n (%) | 8 (100) |
| Frequency of outpatient visits: |  |
| At least once a month: n (%) | 1 (12.5) |
| At least every 3 months: n (%) | 4 (50) |
| At least every 6 months: n (%) | 2 (25) |
| At least every 12 months: n (%) | 1 (12.5) |
| History of mental health condition diagnosed by a health professional, n (%) | 1 (12.5%) |
| Anxiety | 1 (12.5%) |
| Main language spoken at home: English, n (%) | 8 (100) |
| Child clinic attendance |  |
| Diabetes, n (%) | 3 (37.5) |
| Neuromuscular, n (%) | 5 (62.5) |
| SEIFA disadvantage category | — |
| Low (deciles 1–3), n (%) | 4 (50) |
| Mid (deciles 4–7), n (%) | 1 (12.5) |
| High (deciles 8–10), n (%) | 3 (37.5) |
| **Clinicians** | **n = 8** |
| Profession, n (%) | 8 (100) |
| Allied health | 1 (12.5) |
| Nursing | 1 (12.5) |
| Nurse consultant | 2 (25) |
| Paediatrics | 4 (50) |
| Years of practice: mean (range) | 20.5 (10-35) |
| Gender: female, n (%) | 6 (75) |
| Clinic: diabetes, n(%) | 5 (62.5) |

SEIFA, Socio-Economic Indexes for Areas.

**Table 2.**
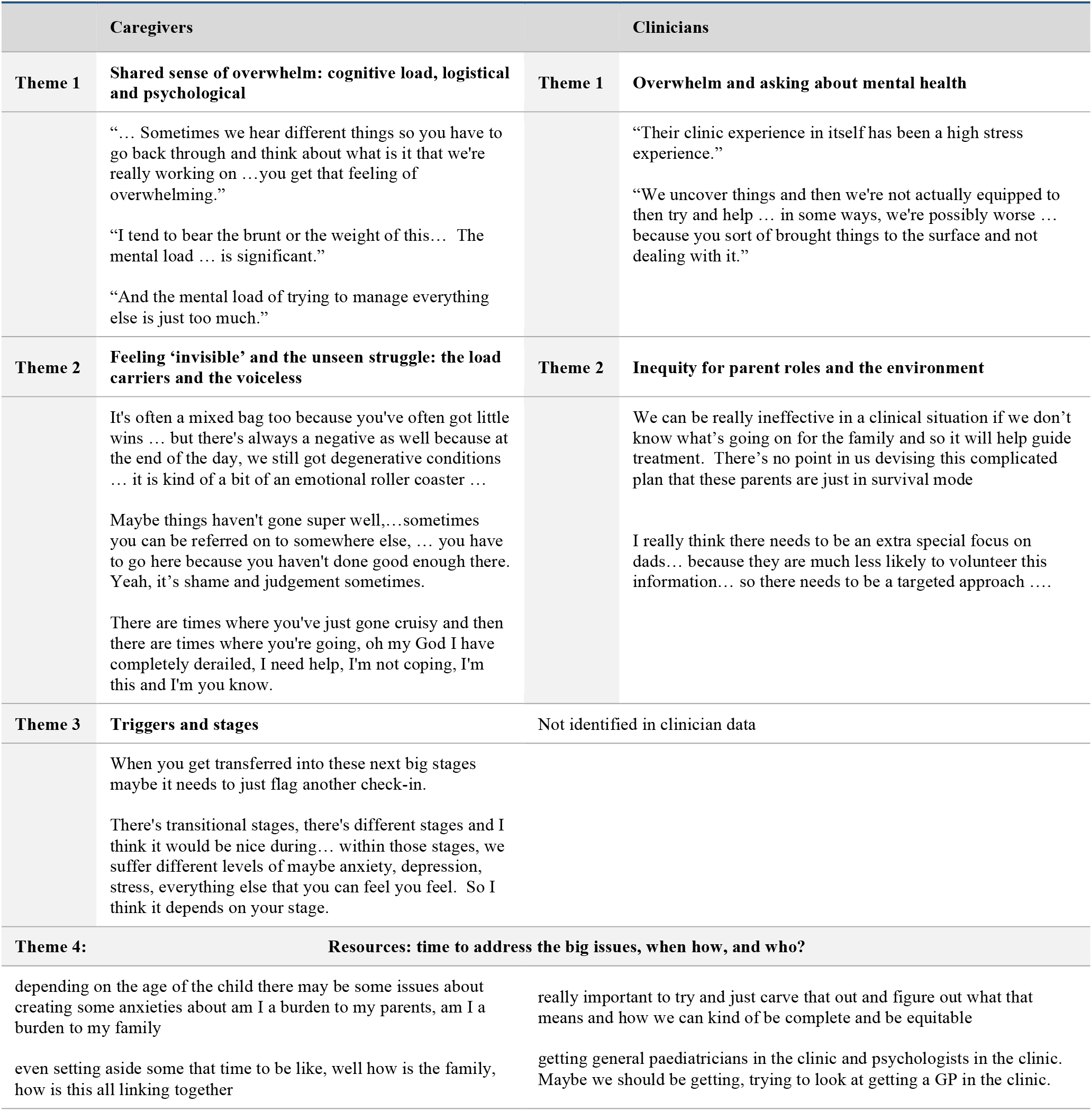
Domain 1: Addressing caregiver mental health. *This domain is about recognising the impact of chronic medical conditions on families and the responsibility of health care professionals to have the conversation*

**Table 3.** Domain 2: Housing the mental health conversation. *This domain is about how to approach screening caregivers and pathways to provide supports for caregivers who are ‘drowning’*

| Caregivers | Clinicians |
| --- | --- |
| **Theme 1: Preferences and reactions to screening questions and delivery** |  |
| DASS 21: "It's going to be a little bit more subtle and a little bit more easier for people to be honest."<br>"One's asking a lot more questions and does that feel more intrusive, more burdensome, do people feel like, I've got to get through more questions. But then I think it's... even as I was looking at them, I can see how asking more questions and in a slightly different way and adding in the physiological stuff might get a bit more of a rounded perspective and capture people."<br>"I think a week before your appointment would be good ...when (child) goes into [condition] clinic ... I normally just sit there." | "Of having a couple of different ways to do that might be helpful, whether it's, some people can complete it in advance but others you can give them the option of while they're sitting in the waiting room before their appointment filling something out."<br>"...In advance of them attending a clinic appointment... so we see our guys six monthly, if it was something that you offered in some way ... and it becomes an opt-in thing." |
| **Theme 2: Care pathways for continuing the conversation, and supporting caregivers** |  |
| "It would depend on how it was flagged because I think there's probably three different groups. There's probably one that just needs acknowledgement and validation that the concerns are real and then that may be all that they need to hear. And then there might be people who need kind of practical support as to... it might be that that's got to do with care issues."<br>"The peer support, because even today, listening to other people's experiences with appointments and that kind of thing too, it's like that's the first time I've ever heard that, and it does often feel like it's a lonely journey when you don't know other people that are in the same situation." | "Having a response that comes back to the parent and says you're scored in these areas and this is what we recommend... you'd want to tailor it slightly differently depending on the clinic because some of the resources that we might suggest in terms of parental peer support groups and things would be slightly different."<br>"Both written and verbal feedback and if we were getting something to take away, I think sending them to a website is not going to work. They're going to need something in their hand to take somewhere else."<br>"I don't think there's any point in burdening parents with yet another piece of paper if it just says 'normal'. But if you're making a recommendation that they then go to their GP to discuss further or ... to sidestep into our psychologist ...then some sort of written information for that purpose would be important." |

**Figure 1.**
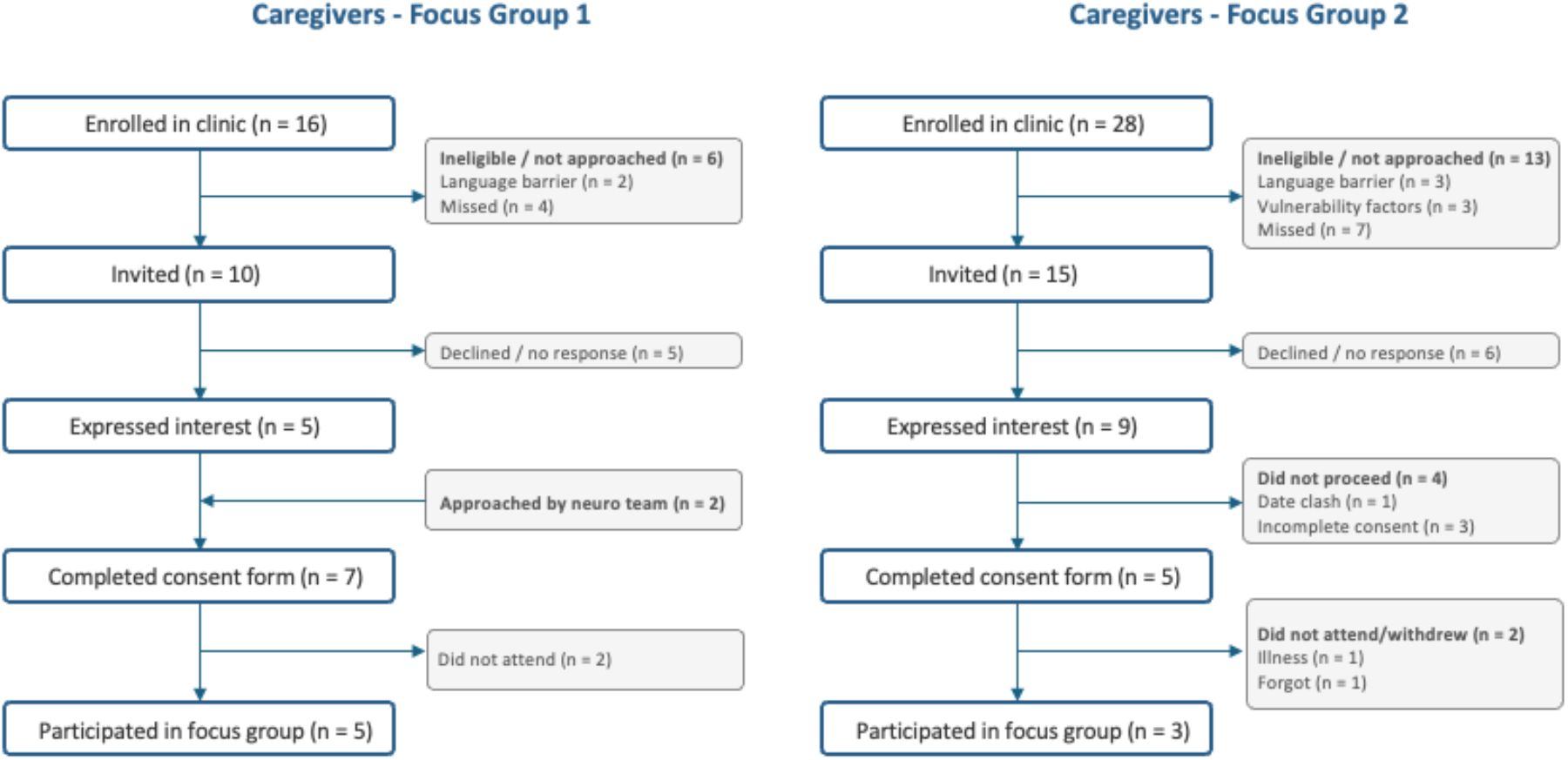
Caregiver focus group recruitment and participation. This flowchart shows recruitment and participation for caregivers across the two focus groups.

**Figure 2.**
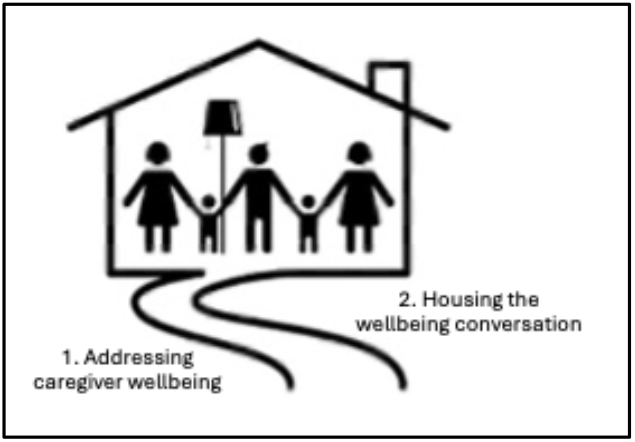
Visual representation of the domains identified through focus group discussion of caregivers and clinicians on caregiver mental health screening in paediatric outpatient clinics. The domains are Addressing Caregiver Mental Health and Housing the Mental Health Conversation.

## Domains and themes

### Domain 1. Addressing the wellbeing conversation

#### Shared sense of overwhelm

Caregivers described a cumulative sense of overwhelm arising from the psychological, logistical, and cognitive demands of managing their child’s condition. This burden extended beyond clinic visits to include ongoing “homework” and the emotional toll of navigating appointments and expectations. “One of the things that I really struggle with is often the amount of homework, or things that come out of an appointment.” For some, this resulted in disengagement from care processes, reflecting both emotional fatigue and feelings of judgement: “I just didn’t feel up to taking my [child] into the appointments; I was sick of getting told off, I’m sick of feeling bad about it.”

Clinicians described a sense of overwhelm in the mental health conversation centring on tension between their ability to respond to caregiver mental health concerns due to outward pressures: “Thinking of my 20 minute …consultation and then all this stuff comes out and just with the resourcing … our patients become more complex, more numerous, but our resources have gone down.*”*

#### The unseen struggle and inequitable roles

Caregivers reflected on the invisibility of many of the emotions they carried and described wanting to protect their children from any additional distress that may be caused talking about the caregiver’s own wellbeing: “…Feel bad or guilty and not going to cause more shame to you as a parent.” Caregiver roles were also strongly gendered, with mothers often positioned as holding the family together, while fathers’ experiences were perceived as less visible and less frequently acknowledged: “I think it’s really important for mums to have the support…we are like the glue…and we have to try and keep it all together.” They highlighted how things differed from the roles of fathers: “Dads … have different things that they’re dealing with these diagnoses as well…. I think it would be nice to be acknowledged… by the team.”

Clinicians reflected on inequities in caregiver visibility, particularly regarding fathers, who were often present at diagnosis but less visible in ongoing cares: “I always worry about the dads I see at diagnosis but never again.” This highlighted assumptions around caregiving roles within paediatric care. Clinicians also described the tension between validating caregiver experiences and protecting children from emotional distress: “Some kids that can’t do that. It would be too much for them … It’s not our place to reveal it to them…”

#### Triggers and stages

Caregivers identified specific stages in the illness trajectory as points of increased emotional burden, particularly at diagnosis as important periods when check-ins would be most valuable: “When… first …diagnosed, just to have that someone from the … clinic that understands… or give you some advice as to where you can go to get further help …and keep it together for the family and keep it … for everyone, when in actual fact most of the time… I feel like I’m drowning.”

#### Resources – time to address the big issues, when, how and who?

Caregivers highlighted structural resource issues as barriers to engaging in mental health conversation within outpatient are, including limited time: “We have very short appointments, so the time with the doctors is very brief.” There was a strong preference for a dedicated contact person who could support ongoing conversations: “I wonder if there is room or funding …for an overarching person we can check in with.” Having a dedicated person was built on the need for trust in these conversations, as caregiver described a tendency to withhold concerns in the absence of an establish relationship: “If you haven’t got that trust.. I say ‘fine’ because…I don’t want to be anything other than fine.”

Clinicians highlighted similar systemic barriers around time and lack of clear pathways to support with suggestions for a dedicated wellbeing role and the importance of long-term relationships: “Maybe there needs to be someone else who’s flagged to check in with these families before or after they see us” or “If we were talking ideal world, it’d be really cool if the Children’s itself drove… like a community of practice where the specialty mental health area was the parents with the lifelong relationship with the hospital.” Clinicians also emphasised that any approach would need to be thoughtfully implemented to avoid being tokenistic: “If we’re having these conversations, then it needs to not feel like a tick box exercise.”

### Domain 2. Housing the wellbeing conversation

#### Preferences and reactions to screening questions and delivery

Caregivers expressed a clear preference for the DASS-21 with its language capturing a more personal reflection of their lived experience: “When I saw … [K10] I thought, ‘Oh, that is so cold,’… I do prefer …[DASS-21] because I just feel like it’s a bit more personal, human … more relatable…” The more specific nature of the questions appeared to prompt reflection on feelings that may otherwise go unrecognised in the context of ongoing caregiving demands: “When you’re in survival mode and you’re just go, go, go, then you’re not necessarily considering, do I feel worthless or what’s going on here?” Preferences for screening administration timing varied, but many caregivers favoured completing screening prior to appointments: “Sometimes you might need it the week before your appointment, a check-in to see how you’re going, sometimes at your appointment.”

Clinicians echoed the emotional implications of the questions in the DASS 21: “They make me feel a bit sad, just reading them…what we’re asking people, it’s quite intense.*”* They also highlighted the importance of accessibility and clarity, particularly when working with diverse populations: “We work with people, all different cultural backgrounds… different intellectual abilities, and you might not know what “hopeless” means… [DASS-21] really uses clear examples, and it makes you reflect on the way you’ve been feeling.” Similarly to caregivers, clinicians felt it was most useful offering the questionnaire prior to the appointment: “So you have a chance to think… almost like an offer to initiate the conversation about it.”

#### Care pathways for continuing the conversation and supporting caregivers

Caregivers emphasised the importance of accessible, supportive follow-up pathways and opportunities for personalised engagement seen as validating: “I think a phone call would be nice

…acknowledge how you are feeling and what you’re going through emotionally.” Caregivers also raised concerns about how sensitive information would be documented and shared, particularly in relation to their child’s medical record: “How does that get documented, or will I see that on my [child’s] letter… and what’s going to happen with that.” Suggested support pathways included both condition-specific, with an emphasis on clarity and practical guidance and to minimise additional tasks: “Some actual direction, or something useful that says, ‘This is what you can do now.’” *and*”…In a way that is not another hurdle you have to jump through…”

Clinicians similarly emphasised the need for practical and accessible support pathways visually presented for maximum benefit: “…beneficial to have it written so that they can take it away and digest it… could use that information to provide to their GP [general practitioner].” Clinicians also felt the value of having access to a list of professionals with expertise: “Development of a database of psychologists … that have experience with dealing with families that experience chronic illness.” Concerns around documentation were also prominent, particularly in relation to confidentiality and the distinction between caregiver and child information: “you’re there for your child … but then once you’re in there as a parent talking about your own private things, it’s a bit muddied.”

## Discussion

With research already showing that caregivers of children with CCs welcome screening on mental health in clinical appointments, evidence on how such screening can be delivered and supported – informed by end-users – has been limited. This study addresses that gap, providing new evidence on caregiver and clinician perspectives on how mental health screening can be embedded into outpatient paediatric care. Two domains emerged from the qualitative analysis that directly addressed these aims: *addressing the wellbeing conversation* and *housing the wellbeing conversation*. Importantly, caregivers demonstrated specific and, at times, unexpected guidance on their preferences for screening measures, agency on feedback delivery and record-keeping, and transition points that would most benefit from mental health supports. In the discussion below, we explore the themes within these domains – including the sense of overwhelm and cognitive load, the load carriers and trigger points for screening – before turning to caregiver and clinician preferences for the DASS-21 questionnaire, and caregiver agency in feedback delivery, confidentiality, and accessible support pathways.

The domain of addressing caregiver mental health captured the cumulative emotional and logistical demands of caregiving as ‘drowning’ and the associated sense of overwhelm. The profound impact of being a caregiver and its potential impact on wellbeing is seemingly unrecognised or not acknowledged. This is reflected in a previous study in an Australian hospital where only 7% of 162 families attending outpatient appointments were asked about caregiver mental health.^32^ Clinicians recognised the importance of these conversations but described constraints related to time, resources and uncertainty about how to respond to complex disclosures. This is also reflected in a previous study with clinicians who identified issues discomfort in being able to manage caregiver mental health conversations.^33^ Delivering holistic care for children with CCs and their caregivers requires that clinicians have the capacity and support to broadly canvass these conversations in clinic appointments.

Caregivers highlighted the emotional labour associated with caregiving roles, including gendered expectations, with mothers often positioned as primary “load carriers”. Both mothers and fathers identified the need for greater recognition of fathers’ experiences, suggesting support may need to be tailored to different caregiving roles. Clinicians also highlighted this, with reference to fathers possibly needing a different approach. This mirrors existing research showing that fathers of young children (n=154) prefer family GP support over mental health support services, with internet resources favoured over telephone counselling.^34^

Caregivers identified key trigger points in the chronic illness trajectory, particularly at diagnosis and during periods of transition, as times of heightened emotional vulnerability. These stages were described as critical opportunities for pro-active support and wellbeing check-ins. Shock and grief following diagnosis are well-documented,^35,36^ and caregivers report this as the period they are least likely to prioritise support for themselves.^36^ Recognition of these trigger points such as changes in disease progression and life transitions may provide an opportunistic timepoint for screening for mental health or check-in with the caregivers.

Caregivers and clinicians overwhelmingly preferred the longer DASS-21 questionnaire and described its questions as more ‘relatable.’ Clinicians prioritised cross-cultural equity in instrument selection, supported by recent evidence of the DASS-21’s reliability and sensitivity across four countries (Germany, Ghana, India and New Zealand).^37^ Both caregivers and clinicians felt that any screening questionnaire should be offered before appointments and then on arrival. Any programs around screening mental health should consider the need for flexibility in delivering questionnaires and at times that suit the specific patient cohort.

Caregiver input into measure preference, feedback delivery and private-recording keeping appears to be unique in existing literature on caregiver mental health screening. While studies have explored systematic mental health screening with caregivers of children with CCs,^38,39^ caregivers do not appear to have been directly consulted on implementation preferences. This is particularly important as any successful screening programs must be acceptable to those with lived experience. A recent systematic review found that involving end-users in the mental health program design helped overcome implementation challenges such as retention, providing protected spaces for discussions, and integrating both medical perspectives and individual experiences.^40^

Caregivers and clinicians were strongly aligned on the importance for clear, practical and accessible support pathways and caregiver autonomy on information delivery and records. Suggestions for support pathways should be useful and tangible. These reflections closely mirror the challenges identified by CF teams who implemented mental health screening programs, particularly separating caregiver records and referral pathways.^23^ These findings further reinforce the need for system-level approaches that integrate caregiver mental health support into routine care, rather than relying solely on individual clinician capacity.

### Strengths and limitations

Strengths include using focus groups allowing sensitive discussion to occur in a more relaxed setting than individual interviews.^41^ Combining caregivers and clinicians reflects evidence that mixed groups yield richer feedback,^42^ while breakout rooms in the second part minimised the potential for power imbalance.^43^ Limitations include a lack of fathers in the caregiver group, and the smaller number of participants in one focus group. Notably, the limited participation of fathers may itself reflect the gendered caregiving dynamics identified in the findings. Findings may also only be transferable to the two condition cohorts, rather than a broader group with other CCs.

## Conclusion

Caregivers and clinicians are both supportive of conversations about caregiver mental health, particularly at specific points on the illness journey. This study uniquely contributes by directly consulting caregivers on preferences regarding screening and feedback delivery, record-keeping and support services. These findings have informed the design of a feasibility trial grounded in caregiver and clinician perspectives.

## Data Availability

All data produced in the present study are available upon reasonable request to the authors.

## Abbreviations

## Acknowledgments

Thank you to Nyanhial Yang for assisting facilitating at a focus group and providing reflective notes. Thank you to caregivers and health professional who took the time to provide their insights, experience and knowledge in this space.

## References

1. Chronic disease Overview. Australian Institute of Health and Welfare. June 17, 2024. Accessed May 8, 2026. https://www.aihw.gov.au/reports-data/health-conditions-disability-deaths/chronic-disease/overview

2. Wisk LE, Sharma N. Prevalence and Trends in Pediatric-Onset Chronic Conditions in the United States, 1999-2018. Acad Pediatr. 2025;25(4):102810. doi:10.1016/j.acap.2025.102810

3. Forrest CB, Koenigsberg LJ, Eddy Harvey F, Maltenfort MG, Halfon N. Trends in US Children’s Mortality, Chronic Conditions, Obesity, Functional Status, and Symptoms. JAMA. 2025;334(6):509–516. doi:10.1001/jama.2025.9855

4. Australia’s children, Chronic conditions and burden of disease. Australian Institute of Health and Welfare. February 25, 2022. Accessed April 9, 2026. https://www.aihw.gov.au/reports/children-youth/australias-children/contents/health/chronic-conditions-burden-disease

5. Aier A, Pais P, Raman V. Psychological aspects in children and parents of children with chronic kidney disease and their families. Clin Exp Pediatr. 2021;65(5):222–229. doi:10.3345/cep.2021.01004

6. Silver EJ, Westbrook LE, Stein REK. Relationship of Parental Psychological Distress to Consequences of Chronic Health Conditions in Children. J Pediatr Psychol. 1998;23(1):5–15. doi:10.1093/jpepsy/23.1.5

7. Cousino MK, Hazen RA. Parenting stress among caregivers of children with chronic illness: a systematic review. J Pediatr Psychol. 2013;38(8):809–828. doi:10.1093/jpepsy/jst049

8. Fairfax A, Brehaut J, Colman I, et al. A systematic review of the association between coping strategies and quality of life among caregivers of children with chronic illness and/or disability. BMC Pediatr. 2019;19(1):215. doi:10.1186/s12887-019-1587-3

9. Cohn LN, Pechlivanoglou P, Lee Y, et al. Health Outcomes of Parents of Children with Chronic Illness: A Systematic Review and Meta-Analysis. J Pediatr. 2020;218:166–177.e2. doi:10.1016/j.jpeds.2019.10.068

10. Pinquart M. Posttraumatic Stress Symptoms and Disorders in Parents of Children and Adolescents With Chronic Physical Illnesses: A Meta-Analysis. J Trauma Stress. 2019;32(1):88–96. doi:10.1002/jts.22354

11. Quittner AL, Goldbeck L, Abbott J, et al. Prevalence of depression and anxiety in patients with cystic fibrosis and parent caregivers: results of The International Depression Epidemiological Study across nine countries. Thorax. 2014;69(12):1090–1097. doi:10.1136/thoraxjnl-2014-205983

12. Eccleston C, Fisher E, Law E, Bartlett J, Palermo TM. Psychological interventions for parents of children and adolescents with chronic illness. Cochrane Database Syst Rev. 2015;4(4):CD009660. doi:10.1002/14651858.CD009660.pub3

13. Herbert L, Hardy S. Implementation of a Mental Health Screening Program in a Pediatric Tertiary Care Setting. Clin Pediatr (Phila). 2019;58(10):1078–1084. doi:10.1177/0009922819862613

14. Lee BK, Loomba RS. Rates of depression, anxiety, and stress in parents of children with congenital heart disease using the Depression Anxiety Stress Scale. Ann Pediatr Cardiol. 2022;15(4):374–379. doi:10.4103/apc.apc_27_22

15. Diederen K, Haverman L, Grootenhuis MA, Benninga MA, Kindermann A. Parental Distress and Quality of Life in Pediatric Inflammatory Bowel Disease: Implications for the Outpatient Clinic. J Pediatr Gastroenterol Nutr. 2018;66(4):630–636. doi:10.1097/MPG.0000000000001756

16. Verkleij M, De Winter D, Hurley MA, Abbott J. Implementing the International Committee on Mental Health in Cystic Fibrosis (ICMH) guidelines: Screening accuracy and referral-treatment pathways. J Cyst Fibros. 2018;17(6):821–827. doi:10.1016/j.jcf.2018.02.005

17. Graziano S, Spanò B, Majo F, et al. Rates of depression and anxiety in Italian patients with cystic fibrosis and parent caregivers: Implementation of the Mental Health Guidelines. Respir Med. 2020;172:106147. doi:10.1016/j.rmed.2020.106147

18. Graziano S, Ullmann N, Rusciano R, et al. Comparison of mental health in individuals with primary ciliary dyskinesia, cystic fibrosis, and parent caregivers. Respir Med. 2023;207:107095. doi:10.1016/j.rmed.2022.107095

19. Vassilopoulos A, Swartz M, Paranjape S, Slifer KJ. Adolescent and caregiver mental health, pulmonary function, and healthcare utilization in pediatric cystic fibrosis. Child Health Care. 2022;51(2):199–212. doi:10.1080/02739615.2021.2002695

20. Warnink-Kavelaars J, Van Oers HA, Haverman L, et al. Parenting a child with Marfan syndrome: Distress and everyday problems. Am J Med Genet A. 2021;185(1):50–59. doi:10.1002/ajmg.a.61906

21. Griffiths N, Laing S, Spence K, et al. Mental health screening for parents following surgical neonatal intensive care unit (NICU) discharge. Early Hum Dev. 2024;198:106128. doi:10.1016/j.earlhumdev.2024.106128

22. Coscini N, McMahon G, Schulz M, et al. Screening parents of children with a chronic condition for mental health problems: a systematic review. Arch Dis Child. 2025;110(9):722–728. doi:10.1136/archdischild-2024-328300

23. Quittner AL, Abbott J, Georgiopoulos AM, et al. International Committee on Mental Health in Cystic Fibrosis: Cystic Fibrosis Foundation and European Cystic Fibrosis Society consensus statements for screening and treating depression and anxiety. Thorax. 2016;71(1):26–34. doi:10.1136/thoraxjnl-2015-207488

24. Salley CG, Bakula DM, Driscoll CFB, et al. Call to Action: Screening and Addressing Caregiver Mental Health Needs within all Pediatric Medical Subspecialty Settings as Standard of Care. Clin Pract Pediatr Psychol. 2024;12(3):328–338. doi:10.1037/cpp0000514

25. Engel G. The need for a new medical model: a challenge for biomedicine. Science. Apr 8;196(4286):129–136. doi:10.1126/science.847460

26. Raina P, O’Donnell M, Schwellnus H, et al. Caregiving process and caregiver burden: Conceptual models to guide research and practice. BMC Pediatr. 2004;4(1):1. doi:10.1186/1471-2431-4-1

27. A Tong, Sainsbury, P, Craig, J. Consolidated criteria for reporting qualitative research (COREQ): a 32-item checklist for interviews and focus groups. Int J Qual Health Care. 19(6):349–357. doi:10.1093/intqhc/mzm042

28. Harris PA, Taylor R, Minor BL, et al. The REDCap consortium: Building an international community of software platform partners. J Biomed Inform. 2019;95:103208. doi:10.1016/j.jbi.2019.103208

29. Creswell JW, Creswell JD. Research Design: Qualitative, Quantitative, and Mixed Methods Approaches. Fifth edition. SAGE; 2018.

30. Braun V, Clarke V. Using thematic analysis in psychology. Qual Res Psychol. 2006;3(2):77–101. doi:10.1191/1478088706qp063oa

31. Graneheim UH, Lundman B. Qualitative content analysis in nursing research: concepts, procedures and measures to achieve trustworthiness. Nurse Educ Today. 2004;24(2):105–112. doi:10.1016/j.nedt.2003.10.001

32. Rhodes A, Sciberras E, Oberklaid F, South M, Davies S, Efron D. Unmet developmental, behavioral, and psychosocial needs in children attending pediatric outpatient clinics. J Dev Behav Pediatr JDBP. 2012;33(6):469–478. doi:10.1097/DBP.0b013e31825a70e6

33. Gilson KM, Johnson S, Davis E, et al. Supporting the mental health of mothers of children with a disability: Health professional perceptions of need, role, and challenges. Child Care Health Dev. 2018;44(5):721–729. doi:10.1111/cch.12589

34. Giallo R, Dunning M, Gent A. Attitudinal barriers to help-seeking and preferences for mental health support among Australian fathers. J Reprod Infant Psychol. 2017;35(3):236–247. doi:10.1080/02646838.2017.1298084

35. Parental and child perspectives on adaptation to childhood chronic illness: A qualitative study. doi:10.1177/1359104509338432

36. Moyes A, Abbott T, Baker S, Reid C, Thorne R, Mörelius E. A parent first: Exploring the support needs of parents caring for a child with medical complexity in Australia. J Pediatr Nurs. 2022;67:e48–e57. doi:10.1016/j.pedn.2022.09.018

37. Adu P, Popoola T, Iqbal N, Medvedev ON, Simpson CR. Validating the depression anxiety stress scales (DASS-21) across Germany, Ghana, India, and New Zealand using Rasch methodology. J Affect Disord. 2025;383:363–373. doi:10.1016/j.jad.2025.04.099

38. Popov N, Phoenix M, King G. To screen or not to screen? Exploring the value of parent mental health screening in children’s rehabilitation services. Disabil Rehabil. 2021;43(5):739–745. doi:10.1080/09638288.2019.1635657

39. Tan SW, Tan MY, Chong SC, et al. Exploring Parental Perspectives on Mental Health Screening and Support for Parents of Children With Autism in Singapore: A Descriptive Qualitative Study. Int J Ment Health Nurs. 2026;35(1):e70208. doi:10.1111/inm.70208

40. Veldmeijer L, Terlouw G, Os JV, Dijk OV, Veer JV ‘t, Boonstra N. The Involvement of Service Users and People With Lived Experience in Mental Health Care Innovation Through Design: Systematic Review. JMIR Ment Health. 2023;10(1):e46590. doi:10.2196/46590

41. Liamputtong P, ed. Research Methods in Health: Foundations for Evidence-Based Practice. Third edition. Oxford University Press; 2017.

42. Femdal I, Solbjør M. Equality and differences: group interaction in mixed focus groups of users and professionals discussing power. Soc Health Vulnerability. 2018;9(1):1447193. doi:10.1080/20021518.2018.1447193

43. Donnelly S, Morton S. Creating organisational and practice change through the use of co-operative inquiry groups in healthcare settings. Action Res. 2019;17(4):451–468. doi:10.1177/1476750319855126

